# Tasimelteon (HETLIOZ®) effective in the treatment of Jet Lag Disorder in an 8-hour phase advance; a multicenter, randomized, double-blind, placebo-controlled trial

**DOI:** 10.1101/2020.04.07.20053306

**Authors:** Christos M Polymeropoulos, Michael A Mohrman, Madison S Keefe, Jennifer L Brzezynski, Jingyuan Wang, Lydia S Prokosch, Vasilios M Polymeropoulos, Changfu Xiao, Gunther Birznieks, Mihael H Polymeropoulos

## Abstract

**Background:** Travelers frequently experience Jet Lag Disorder (JLD) symptoms due to misalignment of the circadian rhythm with respect to the new time zone. In the JET8 Study, we assessed the efficacy and safety of tasimelteon (HETLIOZ®) in healthy participants using a laboratory model of JLD induced by an 8-h phase advance of the sleep-wake cycle. We hypothesized that tasimelteon treatment in participants experiencing JLD would cause increased sleep time, increased next-day alertness, and reduced next-day sleepiness.

**Methods:** We undertook a randomized, double-blind, placebo-controlled trial in 12 US clinical research sleep centers. We screened healthy adults ages 18-73 years, who were eligible for the randomization phase of JET8 if they typically went to bed between 21:00 and 01:00, slept between 7-9 hours each night, and slept at a consistent bedtime. We used block randomization stratified by site to assign participants (1:1) to receive a single oral dose of tasimelteon (20mg) or placebo 30 min before their 8-h phase-advanced bedtime. The primary endpoint was Total Sleep Time in the first 2/3 of the night (TST_2/3_), which was measured by polysomnography during the 8-h sleep episode, and assessed in the intent-to-treat population. The trial is completed and registered with ClinicalTrials.gov, NCT03373201.

**Results:** Between October 16, 2017 and January 17, 2018, we screened 607 healthy participants for JET8, of whom 320 (53%) were assigned to receive tasimelteon (n=160) or placebo (n=160). Tasimelteon treatment resulted in increased TST_2/3_ by 60.3 min (95%CI 44.0 to 76.7, *P*<0.0001) compared to placebo and had a highly significant benefit in secondary endpoints including TST of the whole night, next day alertness, next day sleepiness, and latency to persistent sleep.

**Conclusion:** A single dose of tasimelteon improves symptoms, including sleep and next day functioning in participants, following an 8-h phase advance of the sleep-wake cycle in a laboratory model of JLD simulating eastward trans-meridian travel.

## Introduction

Jet Lag Disorder (JLD), also known as Jet Lag and Circadian Rhythm Sleep-Wake Disorder – Jet Lag Type, is a Circadian Rhythm Sleep-Wake Disorder (CRSWD) characterized by a mismatch between the timing of an individual’s endogenous circadian cycle and the sleep and wake patterns required following a rapid change in time zone. After travelling across two or more time zones, an individual’s endogenous circadian clocks become misaligned with the destination’s local time zone.^1,2^ The mismatch between a person’s intrinsic circadian cycle and their extrinsic 24-h light-dark cycle may cause them to experience symptoms of JLD due to a misaligned circadian rhythm with respect to the new time zone. The essential features of JLD, according to the International Classification of Sleep Disorders Third Edition, are night-time insomnia and daytime sleepiness.^1^ Other symptoms that may be associated with JLD include impairment of daytime functioning and malaise.^3^ Excessive daytime sleepiness and circadian misalignment can lead to impaired concentration, attention, performance, and alertness.^4^ JLD can also be accompanied by gastrointestinal symptoms, light-headedness, headache, fatigue, general malaise, decreased appetite, indigestion, and menstrual symptoms in women, but are not cardinal features of the disorder.^5^

The endogenous circadian clock drives an intrinsic 24-h rhythm in humans that regulates hormone levels, body temperature, metabolism, and the sleep-wake cycle. The circadian timing system (CTS) is composed of a hypothalamic circadian pacemaker located in the suprachiasmatic nuclei (SCN), an array of SCN outputs, and a system of molecular clocks in peripheral tissues.^6^ The light-dark cycle is the major environmental time cue and most powerful synchronizer (“zeitgeber”) of the CTS to the Earth’s 24-h day. Light sensed by intrinsically photosensitive Retinal Ganglion Cells (ipRGCs) provides information on the daily light-dark cycle to the SCN via the retinohypothalamic tract (RHT). Through this input, the SCN is synchronized (entrained) to a 24-h light-dark cycle. The SCN also functions as the master body clock by entraining the body’s peripheral clocks to a 24-h rhythm through endocrine, neuronal, and physiological signals, which regulate cyclic levels of melatonin, cortisol, and core body temperature.^7,8^ Through the synchronization of peripheral clocks, the SCN governs the optimal timing of key physiological processes, including those of the cardiovascular system, metabolism, immune regulation, rest activity, and the sleep-wake cycle.^9-11^

Sleep disruption is the primary complaint of JLD, especially when travelling in an eastward direction, which results in a phase advance of sleep timing and circadian rhythmicity.^5,12^ Following eastward travel, the first two thirds of the night is usually very disrupted, as that portion of the night overlaps with the traveler’s circadian timing. Due to the phase advance associated with eastward flights across 3-8 time zones, travelers have difficulty going to sleep at the new bedtime, whereas after westward flights across 3-8 time zones, travelers have difficulty remaining awake in the evening and have early morning awakenings in the new time zone.^13,14^ Additionally, depending on the number of hours of the phase advance and speed to physiologically adjust to the new time zone, the main effect can be on sleep latency and may only affect the first night or two of the trip.^15^ For instance, west coast to east coast US travel results in a 3-h phase advance, which is equivalent to being required to sleep three hours before an individual’s normal bedtime. This would result in difficulty falling asleep, but after the first three hours, the individual would be attempting to sleep during their typical sleep cycle. Additionally, for this phase advance, the effect may only be experienced on the first night, with adaptation to the new time zone on subsequent nights. We have shown this propensity for adaptation in a previous study.^15^

The hormone melatonin has a robust endogenous circadian secretory profile that peaks during night hours. Aberrant melatonin rhythms are associated with CRSWD including Non-24-Hour Sleep-Wake Disorder (Non-24), Delayed Sleep-Wake Phase Disorder, and JLD. Two types of G-protein-coupled receptors for melatonin, MT_1_ and MT_2_, are located in the SCN. Melatonin exerts sleep-promoting effects, although the exact mechanism is not clear. It has been suggested that the sleep-promoting effects may result from melatonin’s ability to induce hypothermia.^16^

Tasimelteon is a Dual Melatonin Receptor Agonist (DMRA) that has been shown to regulate the timing of melatonin secretion when administered to participants in advance of the usual dim light onset of melatonin.^17^ We have previously shown that tasimelteon can entrain the circadian clock of individuals who are not entrained to the 24-h day and have Non-24 by administering tasimelteon every night 30 minutes before the target bedtime.^17^ We have further shown that administration of tasimelteon advances the circadian clock during a 5-h phase advance.^15^ It has also been reported that exogenous melatonin administration can entrain and shift the circadian clock in humans.^18-20^ Tasimelteon has not previously been tested to phase advance individuals more than 5 hours, and there are no phase response curve data available for tasimelteon. However, phase response curve data for melatonin suggests that the maximal resetting effect of melatonin for inducing a phase advance occurs at about 3-h before dim light melatonin onset (DLMO) timing (about 5-h before the start of the scheduled sleep episode) and can advance the endogenous circadian phase by about 1-h after one night.^20^ Further, it is described that phase advances of longer than 7 hours can induce an “antidromic” response, wherein the individual will start delaying to the new phase instead of advancing (for instance, delaying 16-h instead of advancing by 8-h).^12^ It is therefore of great interest to conduct a clinical study to determine the effects of administering a melatonin receptor agonist following a phase advance in sleep timing of more than 5-h.

Here we present the results of the JET8 study, a phase III clinical trial to determine the efficacy and safety of a single oral dose of tasimelteon in healthy participants in a laboratory simulation of JLD associated with an 8-h phase advance of sleep-wake timing.

## Methods

### Study design

The study used a randomized, double-blind, placebo-controlled, parallel design and was approved by institutional review boards, either Chesapeake IRB or BioMed IRB, at all sleep centers. Screening consisted of a clinical site visit to evaluate eligibility followed by a screening interval lasting from one to four weeks. Evaluation, which consisted of a clinical site overnight stay, followed successful completion of the screening procedure (Figure 2). Additionally, the end of Daylight Saving Time (DST) occurred during the study. Study sites were instructed not to schedule any participants for evaluation for the day before, or the week following, November 4, 2017. This was done to allow participants to naturally adjust to the 1-h phase delay induced by DST. Twelve US study sites with single-bed suites were utilized for the evaluation visit. Suites were free of time cues such as light and sound.

**Figure 2.**
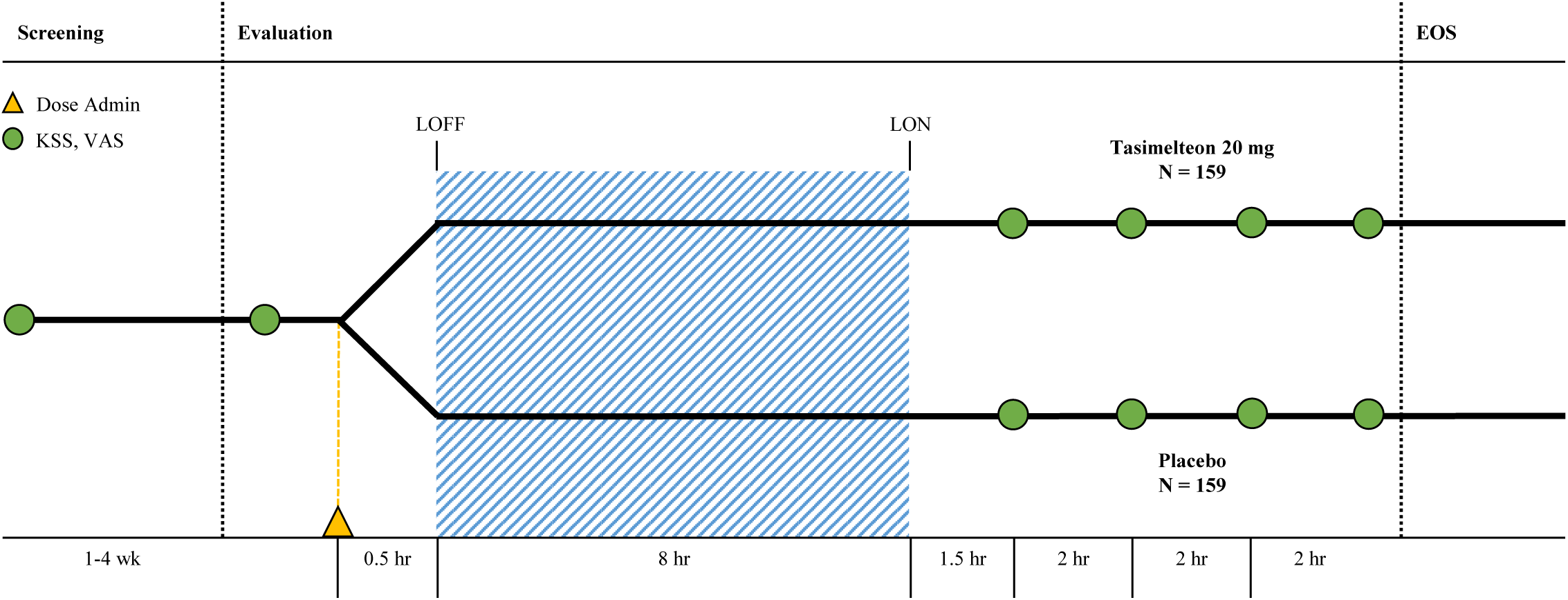
Study Design. The study consisted of a 1-4 week screening period followed by an evaluation visit where patients were dosed with tasimelteon 20mg or placebo 30-minutes prior to their pre-determined 8-hour phase advance bedtime. Polysomnography was performed during the 8-hour sleeping period. Karolinska Sleepiness Scale (KSS) and Visual Analogue Scale (VAS) assessments were completed once during screening, once before the phase advance, and four times after waking.

### Participants

Participants recruited were men and women aged between 18 and 73 years, in good health (determined by medical and psychiatric history, physical examination, electrocardiography, serum chemistry, hematology, urinalysis, and urine toxicology) and without major sleep disorders (assessed with validated questionnaires). Study participants provided written informed consent before any screening procedures began.

At the inpatient screening visit, participants self-reported their habitual bedtime and self-determined target bedtime. Participants were excluded at screening if their habitual and target bedtimes were not between 21:00 and 01:00, or if their average habitual total sleep window were less than 7-h, or more than 9-h. After completion of the screening visit, participants were recruited to maintain a consistent sleep-wake schedule for one to four weeks. Participants were instructed to adhere to their self-selected target bedtime for the duration of screening. Participants were eligible for randomization if they met their target bedtime, ± 30 min, for the three nights prior to the inpatient evaluation visit, and at least five of the seven nights before the evaluation visit. Exclusions also occurred if participants did not have an average sleep episode duration of at least 6.7-h and no more than 9-h for the seven nights before evaluation. For the last three days prior to the inpatient evaluation visit, the participant must have had at least 7-h and no more than 10-h, of self-reported total sleep time for each night.

### Randomization and masking

Randomization was performed through an interactive web response system (IWRS). When each participant arrived at the evaluation visit, the investigator or designee utilized the IWRS system to distribute study medication from the capsule-containing bottle.

Participants were randomly assigned to either tasimelteon (20mg) or placebo, in a 1:1 ratio. All capsules were size 1, opaque, hard gelatin, and the color was dark blue with two white bars printed on both the cap and body of the capsule. Placebo was provided in size and appearance identical to those containing tasimelteon. An unblinded, third-party statistician prepared the randomization scheme. Randomization was stratified by study site.

### Procedures

During screening, assessments taken included the Karolinska Sleepiness Scale (KSS), a Visual Analog Scale (VAS), and a post-sleep questionnaire (PSQ). The KSS and VAS were subjective measures of next day alertness. The KSS queried participants as to how sleepy they felt on a 9-point scale with 1 being extremely awake and 9 being extremely sleepy/fighting to stay awake. The VAS was a self-rated scale to assess sleepiness. Participants marked along a 100 mm line to represent their current state of sleepiness, 0 being very sleepy and 100 being very alert. The PSQ was a self-reported measurement of wake after sleep onset (WASO), sleep latency, total sleep time (TST), number of nocturnal awakenings, and overall sleep quality. The overall sleep quality was reported on a scale from 1 to 5, corresponding to “poor”, “fair”, “average”, “good”, or “excellent”.

Daily electronic diaries and wrist actigraphy watches were used to ensure compliance with eligibility parameters during outpatient screening. These eligibility parameters were used to ensure that the participants were sleep satiated in the days preceding the inpatient overnight sleep phase advance.

For the 8-h sleep-wake phase advance, the participants’ bedtime was advanced by 8-h compared to the target bedtime established during screening (Figure 3). Single oral dose administration of medication, tasimelteon or placebo, occurred 30 min (± 5 min) prior to lights off. Tasimelteon and placebo were manufactured by Patheon Pharmaceuticals Inc. in Cincinnati, Ohio, US. Participants’ sleep was monitored by polysomnography (PSG), and centrally scored in 30-s epochs. Polysomnographic data were analyzed for the single inpatient 8-h sleep period during the evaluation visit.

**Figure 3.**
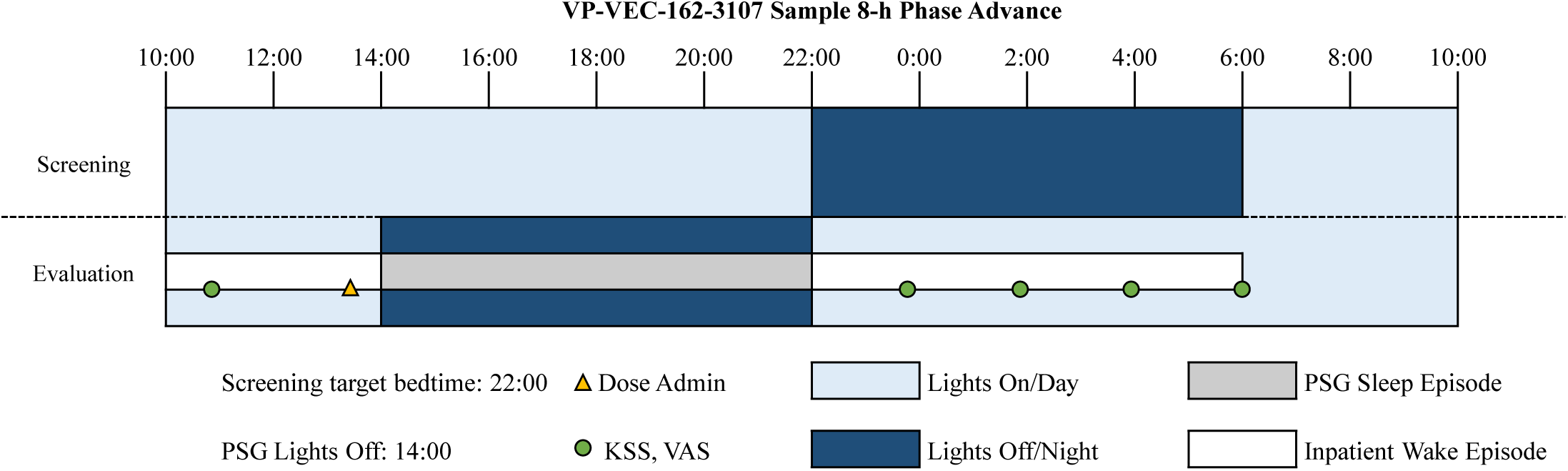
Raster Plot of Phase Advance Protocol. An example phase advance for a participant with a screening target bedtime of 22:00 and an 8-hour phase advanced bedtime of 14:00. The 8-hour phase advanced bedtime was determined for each patient based on their screening target bedtime.

During evaluation, assessments taken included the KSS, VAS, and PSQ. The KSS and VAS were each administered four times following the dose administration after wake-up. The first KSS and VAS were conducted 90 min (± 15 min) after “lights on”, and the following three KSS and VAS were conducted once every 2-h, over the following 6-h. The PSQ was administered once, approximately 30 min (± 15 min) after lights on, following the 8-h phase advance.

### Outcomes

The objective polysomnographic primary outcome measure was total sleep time in the first two-thirds of the night (TST_2/3_) following an 8-h phase advance of the sleep episode. TST_2/3_ was selected as the primary endpoint for multiple reasons. The first two-thirds of the night are maximally overlapped with the previous circadian day and therefore of most interest to understand sleep during a circadian adverse time. Further, many individuals habitually sleep less than 8 hours per night and therefore wakefulness in the third third of an 8-h interval may represent the normal morning after a sufficient night of sleep.

Secondary objective outcomes measured by PSG included TST, WASO, and latency to persistent sleep (LPS) for additional time intervals of the night. WASO was defined as the time spent awake in the interval between onset of persistent sleep and lights on. LPS was defined as the length of time elapsed between lights off and onset of persistent sleep. Subjective secondary outcome measures included the next day residual effects of tasimelteon, as measured by individual and average KSS scores, individual and average VAS scores, and the PSQ. Average Night 1 KSS and Average Night 1 VAS combined all four KSS and VAS assessments collected after dose administration. These primary and secondary outcomes were centrally assessed across all study centers.

Safety assessments included regular monitoring and recording of adverse events; monitoring of hematology, serum chemistry, and urinalysis values; monitoring of vital signs; performance of physical examinations; and performance of electrocardiograms. Safety assessments were performed at the inpatient screening and evaluation visits, and any supplementary unscheduled visits.

### Statistical analysis

For hypothesis testing, a sample size of 150 individuals per treatment group was estimated to provide approximately 93% power to detect a 30-min difference in TST_2/3_ following an 8-h phase advance bedtime between the treatment group and placebo based on a two-tailed t test with α=0.05^15^ and a standard deviation (SD) of 75 min, which was estimated from a previous study comparing doses of tasimelteon and placebo in healthy volunteers in a 5-h phase advance model.^15^ Additionally, this sample size provided at least 85% power to detect a difference of 28 min (SD 80) in TST between the treatment group and placebo.

The intent-to-treat (ITT) population was defined as all individuals randomized into the study who received a dose of study drug and had complete PSG data. The difference between treatment groups was summarized by the difference between the least squared means (LS) and the 95% confidence interval (95% CI). Hypotheses tested were declared statistically significant if the calculated *P*-values were ≤ 0.05. All primary and secondary objective outcome measures between treatment groups were analyzed by analysis of variance (ANOVA). The Wilcoxon Rank-Sum test of TST_2/3_ was performed as a sensitivity analysis. All primary and secondary endpoints were pre-specified in the statistical analysis plan. All analyses and tabulations were performed using SAS® version 9.3 or higher. This study is registered with clinicaltrials.gov under NCT03373201.

## Results

Between October 16, 2017 and January 17, 2018 a total of 607 participants were assessed for eligibility in the JET8 study. 320 (52.7%) participants met the inclusion criteria and were enrolled. Following randomization, one participant (0.3%) in the placebo group withdrew consent and one participant (0.3%) in the tasimelteon group discontinued due to a serious adverse event determined to be unrelated to therapy. 318 individuals completed the study and were included in the ITT population analysis (Figure 1).

**Figure 1.**
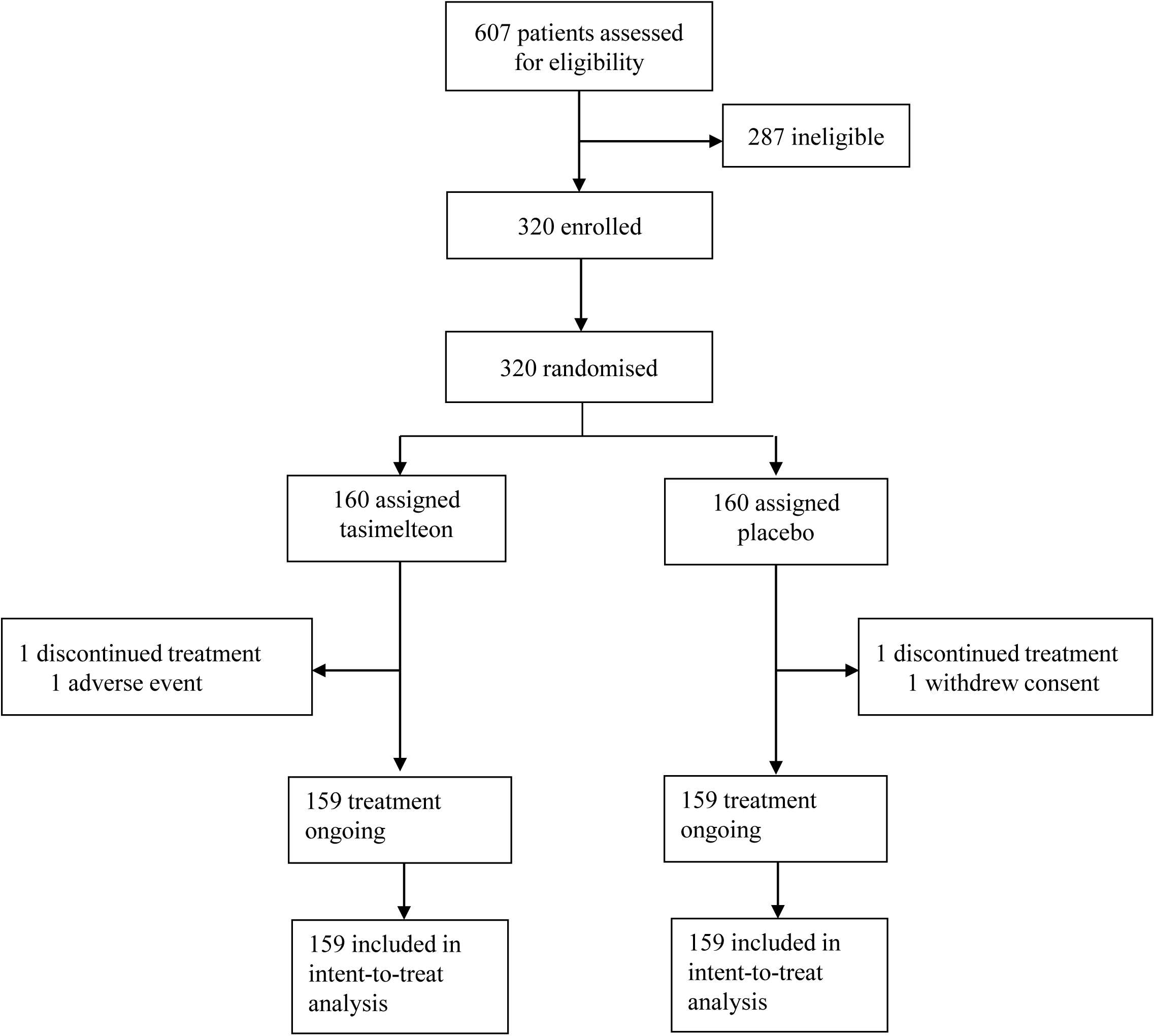
Trial Profile. Number of patients screened, enrolled, randomized, and discontinued for the JET8 study.

Baseline demographics were similar between tasimelteon and placebo groups. Of the 318 ITT participants, 151 (47.5%) were male. The average age was 35.8 (SD 11.92), and average BMI was 25.0 (SD 3.02). The racial breakdown was 191 (60.1%) White, 91 (28.6%) Black or African American, 28 (8.8%) Asian, 0 (0.0%) Native Hawaiian or Other Pacific Islander, 1 (0.3%) American Indian or Alaska Native, and 7 (2.2%) Other (Table 1).

**Table 1:** Baseline Characteristics for the ITT Population.

|  | Tasimelteon 20 mg<br>(N=159) | Placebo<br>(N=159) | Total<br>(N=318) |
| --- | --- | --- | --- |
| Age (year) | 35.8 (11.20) | 35.9 (12.63) | 35.8 (11.92) |
| Sex |  |  |  |
| Male | 73 (45.9%) | 78 (49.1%) | 151 (47.5%) |
| Female | 86 (54.1%) | 81 (50.9%) | 167 (52.5%) |
| Baseline BMI (kg/m<sup>2</sup>) | 25.1 (3.04) | 24.8 (3.00) | 25.0 (3.02) |
| Race |  |  |  |
| White | 94 (59.1%) | 97 (61.0%) | 191 (60.1%) |
| Black or African American | 44 (27.7%) | 47 (29.6%) | 91 (28.6%) |
| Asian | 15 (9.4%) | 13 (8.2%) | 28 (8.8%) |
| Native Hawaiian or Other Pacific Islander | 0 (0.0%) | 0 (0.0%) | 0 (0.0%) |
| American Indian or Alaska Native | 1 (0.6%) | 0 (0.0%) | 1 (0.3%) |
| Other | 5 (3.1%) | 2 (1.3%) | 7 (2.2%) |

Data are n (%), or mean (SD). Baseline is defined as the last non-missing measurement prior to start of study drug. BMI = Body Mass Index.

The primary outcome was TST_2/3_ (0-320 min), following an 8-h phase advance, and was measured by PSG. Tasimelteon treatment resulted in increased TST by 60.3 min (44.0 to 76.7, *P*<0.0001) in the first two-thirds of the night (Table 2).

**Table 2:** Summary of Endpoints for the ITT Population.

|  | Tasimelteon 20 mg<br>(N=159) | Placebo<br>(N=159) | Difference<br>(95% CI) | P-value |
| --- | --- | --- | --- | --- |
| **Primary Endpoint** |  |  |  |  |
| TST<sub>2/3</sub> (min) | 216.4 | 156.1 | 60.3 (44.0 to 76.7) | $P < 0.0001$ |
| **Objective Secondary Endpoints** |  |  |  |  |
| WASO (min) | 144.6 | 219.1 | -74.6 (-94.8 to -54.3) | $P < 0.0001$ |
| LPS (min) | 21.8 | 36.8 | -15.1 (-26.2 to -4.0) | $P = 0.0081$ |
| TST<sub>full</sub> (min) | 315.8 | 230.3 | 85.5 (64.3 to 106.6) | $P < 0.0001$ |
| TST<sub>first third</sub> (min) | 124.6 | 102.4 | 22.2 (14.1 to 30.3) | $P < 0.0001$ |
| TST<sub>second third</sub> (min) | 91.8 | 53.7 | 38.1 (26.7 to 49.5) | $P < 0.0001$ |
| TST<sub>final third</sub> (min) | 99.4 | 74.2 | 25.1 (13.8 to 36.5) | $P < 0.0001$ |
| **Subjective Secondary Endpoints** |  |  |  |  |
| Average Night 1 KSS (pt) | 4.0 | 4.5 | -0.5 (-0.9 to -0.1) | $P = 0.0083$ |
| Average Night 1 VAS (mm) | 60.8 | 54.2 | 6.6 (1.6 to 11.6) | $P = 0.0099$ |
| WASO (min) | 75.3 | 113.5 | -38.2 (-64.2 to -12.2) | $P = 0.0041$ |
| Sleep Latency (min) | 27.0 | 40.9 | -13.9 (-29.8 to 2.0) | $P = 0.0857$ |
| TST (min) | 393.6 | 331.9 | 61.7 (34.5 to 88.9) | $P < 0.0001$ |
| No. Nocturnal Awakenings | 2.6 | 2.8 | -0.3 (-0.7 to 0.2) | $P = 0.2389$ |
| Sleep Quality (pt) | 3.3 | 2.7 | 0.6 (0.3 to 0.9) | $P = 0.0001$ |

Data are average measurements recorded by polysomnography for the primary and objective endpoints and recorded by questionnaire for the subjective endpoints. Tasimelteon treatment resulted in significant improvements in objective and subjective sleep time, wake after sleep onset, latency to persistent sleep, and next day alertness as measured by KSS and VAS.
TST<sub>2/3</sub> = Total Sleep Time in the first 2/3 of the night; TST = Total Sleep Time; WASO = Wake After Sleep Onset; LPS = Latency to Persistent Sleep; KSS = Karolinska Sleepiness Scale; VAS = Visual Analog Scale; No. = Number.

Over the full eight hour sleep episode, TST increased by 85.5 min (64.3 to 106.6, *P*<0.0001) for the tasimelteon group compared to the placebo group. WASO was reduced by 74.6 min (−94.8 to −54.3, *P*<0.0001) for the tasimelteon group compared to the placebo group. LPS was decreased by 15.1 min (−26.2 to −4.0, *P*=0.0081) for the tasimelteon group compared to the placebo group (Table 2). Additionally, for each individual third of the night, average TST was significantly increased in the tasimelteon group compared to the placebo group. Average TST was increased for the tasimelteon group compared to the placebo group in the first third of the night (0-160 min) by 22.2 min (14.1 to 30.3, *P*<0.0001), in the second third of the night (161-320 min) by 38.1 min (26.7 to 49.5, *P*<0.0001), and in the final third of the night (321-480 min) by 25.1 min (13.8 to 36.5, *P*<0.0001) (Table 2). For each hour of the night, average TST was also significantly increased for the tasimelteon group compared to the placebo group (Figure 4).

**Figure 4.**
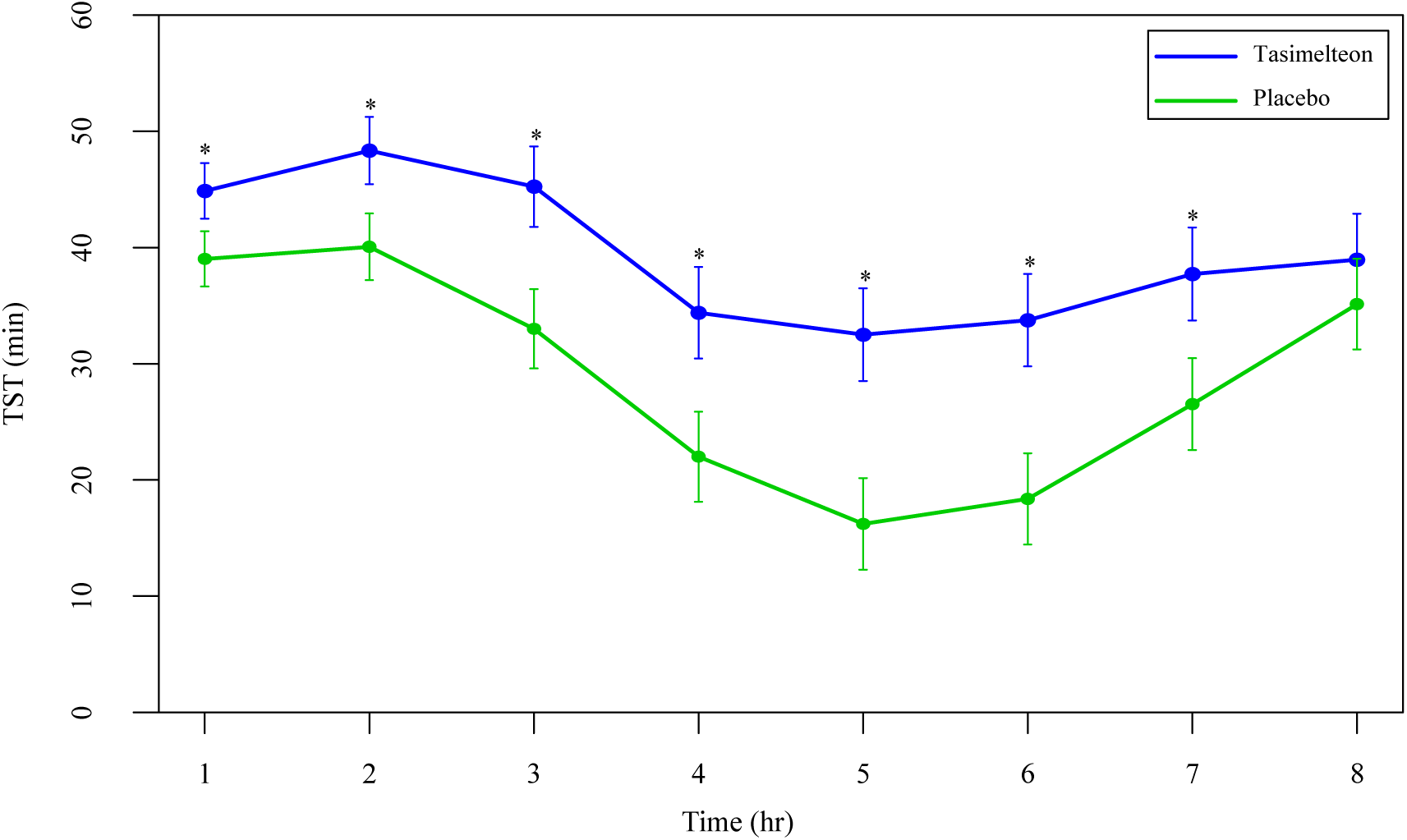
Average Total Sleep Time by Hour. Data are LS Mean (95% CI) for average TST by hour between tasimelteon and placebo. TST during the 8-h phase advance demonstrated a statistically significant increase in the tasimelteon group compared to placebo for hrs 1-7. * *P* < 0.001

Secondary outcome measures of subjective sleep efficacy were assessed by subject reported measures including the KSS, VAS, and PSQ. Average Night 1 KSS was decreased for the tasimelteon group by 0.5 pts (−0.9 to −0.1, *P*=0.0083) compared to the placebo group (Table 2). Average Night 1 VAS was increased for the tasimelteon group by 6.6 mm (1.6 to 11.6, *P*=0.0099) compared to the placebo group (Table 2). KSS and VAS administered at each time point are shown in Figure 5. Subjective measures assessed by the PSQ including WASO, TST, and overall sleep quality were significantly improved for the tasimelteon group. WASO was reduced by 38.2 min (−64.2 to −12.2, *P*=0.0041) for the tasimelteon group compared to the placebo group. TST was increased by 61.7 min (34.5 to 88.9, *P*<0.0001) for the tasimelteon group compared to the placebo group. Overall sleep quality increased by 0.6 pts (0.3 to 0.9, *P*=0.0001) in the tasimelteon group compared to the placebo group. Trends in sleep latency and the number of nocturnal awakenings were also observed (Table 2).

**Figure 5a.**
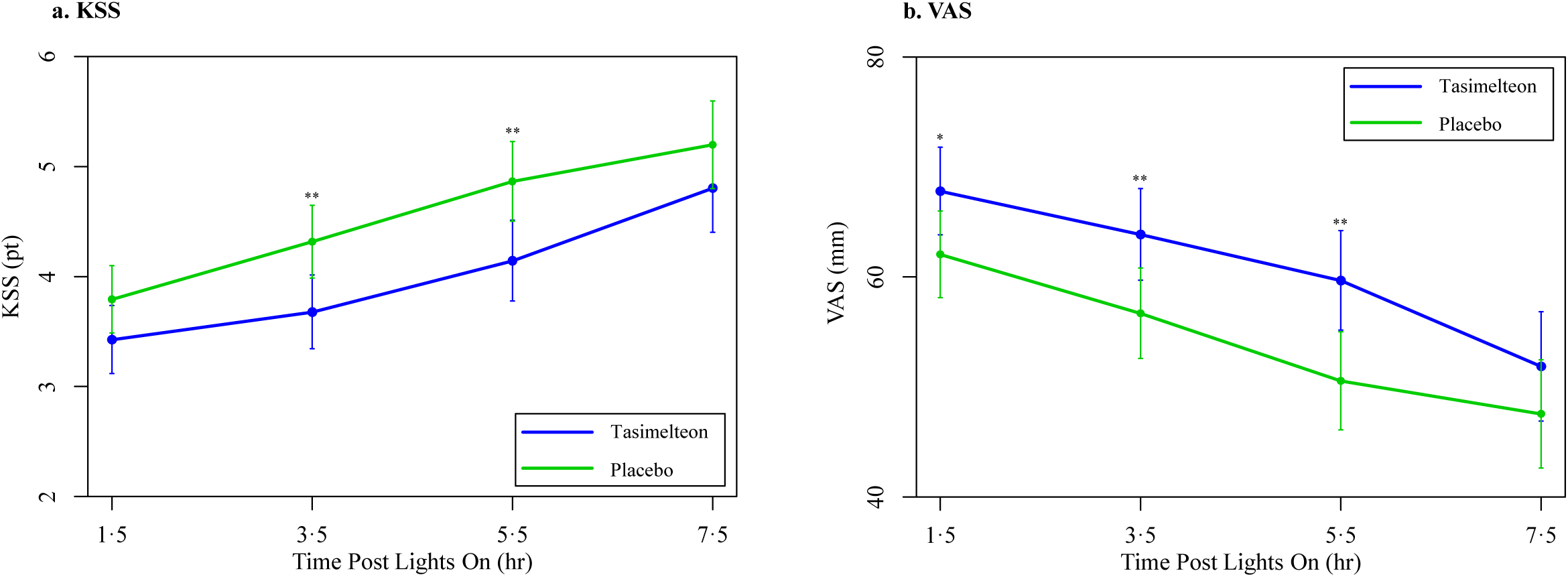
Night 1 KSS & VAS. Data are LS Mean (95% CI) for average Night 1 KSS values between tasimelteon and placebo. KSS values, 1 pt being extremely awake and 9 pts being extremely sleepy/fighting to stay awake, demonstrated a statistically significant decrease following the 8-h phase advance in the tasimelteon group compared to the placebo group for 3.5 and 5.5 hrs post-lights on. ** *P* < 0.01 **Figure 5b.** Data are LS Mean (95% CI) for average Night 1 VAS values between tasimelteon and placebo. VAS values, 0 mm being very sleepy and 100 mm being very alert, demonstrated a statistically significant increase following the 8-h phase advance in the tasimelteon group compared to the placebo group for 1.5, 3.5, and 5.5 hrs post-lights on. ** *P* < 0.01, * *P* < 0.05

The safety analysis included data from 320 participants (tasimelteon n=160, placebo n=160) who received one treatment dose after randomization. Discontinuation due to treatment emergent adverse events was insignificant between treatment groups. The most common treatment emergent adverse event reported in the JET8 study was headache (8[5%] tasimelteon *vs* 4[2.5%] placebo). One serious adverse event was reported but was determined by the Principal Investigator to be unrelated to study drug.

## Discussion

Treatment with 20mg tasimelteon in this study resulted in clinically meaningful improvements in objective and subjective measures of sleep duration and sleep quality for participants exposed to an 8-h phase advance, as travelers experience when flying eastward across eight time zones. Tasimelteon treatment also resulted in increased alertness and decreased sleepiness the day following the phase advance. These improvements are of paramount importance given the safety consequences of reduced alertness and sleepiness on individuals experiencing JLD resulting in occupational hazards and motor vehicle accidents. The symptoms of JLD can be severe in up to a quarter of patients. Further, the fatigue and cognitive effects can result in significant errors when performance is critical, such as when operating machinery or being a member of the flight crew. Although it is well known that sleep deprivation causes significant morbidity and mortality annually as a result of performance impairment, fatigue during times of eastward JLD itself has been implicated as the causative agent of several airplane accidents.^13, 21, 22^

There is currently no treatment for JLD approved by the US Food and Drug Administration (FDA) or the European Medicines Agency (EMA). The American Academy of Sleep Medicine (AASM) recommends the use of light therapy, slowly changing sleep-wake schedule timing in advance of a trip, and the nutritional supplement melatonin, although an appropriate dosage form has not been determined. Caffeine may counteract jet lag-induced sleepiness but may also disrupt night-time sleep.^23^ The AASM acknowledges the use of hypnotic sleeping pills as a rational treatment for JLD; however, they mention hypnotics are not necessary, have risks of adverse effects like global amnesia, and should be used on a short-term basis.^23^ Travelers who follow even the above recommendations continue to report symptoms of JLD.

It is estimated that about two-thirds of people who complete eastward, transmeridian travel across three or more time zones experience at least moderate JLD, and almost one-third experience severe symptoms of JLD.^22,24^ In 2018, 93 million US citizens travelled internationally. Outbound overseas travel from the US totaled 41.8 million travellers.^25^ Of the millions of travelers who cross time zones each year, 80% report disrupted sleep during the scheduled sleep episode in the new time zone.^26^

Although previous studies demonstrated that treatment with oral melatonin after a phase advance may reduce sleep onset latency and improve sleep quality,^18,19^ increasingly studies are focusing on Dual Melatonin Receptor Agonists, as they may have a more stable pharmacokinetic profile, may be more potent, and may produce a greater effect than melatonin.^5,27^ Prior studies with tasimelteon, which has selective agonist activity at the MT_1_ and MT_2_ receptors,^15,28^ have demonstrated that when taken on a fixed time schedule, tasimelteon entrains the circadian clock of blind individuals with Non-24 to the 24-h day.^17^ The master body clock regulates the circadian rhythms of hormones, many aspects of physiology, metabolism, and behavior, including melatonin and cortisol, and synchronizes them with the 24-h day.^6,7^ For this study, we hypothesized that tasimelteon would provide an alternative 24-h time cue necessary to resynchronize the CTS to the newly imposed 24-h day. The robust results observed in this study confirm that participants who took tasimelteon had longer and better quality sleep. Lastly, we found that the morning following tasimelteon dosing, participants experienced fewer symptoms of JLD, as they reported feeling both more alert and less sleepy.

We believe that the underlying mechanism involves tasimelteon replacing the light-dark cycle as the time cue that synchronizes the CTS to the 24-h day. If the CTS of an individual with JLD can be re-synchronized to the new 24-h light-dark cycle, we predict that the master body clock will be able to regulate the secretion of hormones like melatonin and cortisol in phase with the new time zone, thus allowing an individual to feel alert during the day and sleepy at night.

The effects of tasimelteon were observed on the first night of treatment. Participants treated with tasimelteon not only fell asleep faster, but also slept 85.5 min longer than placebo during the 8-h sleep episode. Participants taking tasimelteon reported lower sleepiness scores and higher alertness scores the day after the phase advance, compared to placebo. Participants taking tasimelteon also self-reported falling asleep faster, spending less time awake during the night, longer sleep, and an overall higher sleep quality compared to placebo. In this study, tasimelteon was safe and well tolerated. This clinical trial demonstrates that tasimelteon is effective for treating the symptoms of JLD following a shift in the timing of the sleep-wake cycle, comparable to that required following eastward travel across eight time zones.

Current therapies available for the treatment of JLD have not been validated and do not fundamentally address the underlying circadian dysfunction. These include sedative hypnotics such as zolpidem and eszopiclone, and wake promoting agents such as modafinil. In the largest (N=257) double-blind trial to evaluate the efficacy of melatonin for the treatment of JLD, 0.5 and 5mg of melatonin were reported to be ineffective.^24^ Similarly, treatment of healthy participants with 1, 4, or 8mg of ramelteon failed to improve sleep.^29^ Moreover, 26% of over-the-counter formulations of melatonin, which is weakly regulated by the FDA as a food supplement and not as a drug, have been reported to be contaminated with serotonin, putting users at risk for serotonin syndrome.^30^

Potential limitations of the study include that Jet Lag was induced by an immediate phase advance of the sleep-wake cycle in a sleep clinic, rather than jet travel in the eastward direction. However, the model in the JET8 study helps eliminate confounders unrelated to JLD, which is fundamentally a circadian dysfunction unrelated to the actual travel in a jet plane. Further, this potential limitation was tested in another study with tasimelteon, the JET study, wherein participants flew from the US to Europe. The JET study met its primary endpoint (source: in publication), demonstrating that tasimelteon is effective to treat JLD in a model that uses transatlantic flights. Additionally, JET8 participants were studied and dosed for one night, as opposed to multiple night dosing. However, JLD may affect individuals for only one night, depending on the degree of phase advance, the length of the trip, and the speed at which the individual adapts to the new time zone. Finally, the JET8 study protocol results in a “first night effect”, in which a new sleeping environment induces insomnia. One potential limitation therefore is that a sleep study in a clinic may be at least partially causing first night effect-induced insomnia in addition to phase advance insomnia. However, comparison of the differences in results between this study, which employed an 8-h phase advance, and the JET5 study, which employed a 5-h phase advance, demonstrates that a difference in the number of time zones advanced results in significant differences in JLD symptoms with and without treatment.^15^

The results of the JET8 study demonstrate effectiveness of tasimelteon in treating the symptoms of JLD. The magnitude of the total benefit over placebo is significant and clinically meaningful. The results of the study strongly suggest that tasimelteon may be an effective therapeutic tool in the treatment of individuals with JLD.

## Data Availability

The original contributions presented in the study are included in the article/supplementary files, further inquiries can be directed to the corresponding author/s.

## Contributors

CMP, GB, CX, and MHP contributed to the study concept and design. JW, CX, CMP, and MHP developed the statistical analysis plan. CMP wrote the report in collaboration with MSK, JLB, VMP, LSP, and MAM. CMP revised the report with participation from all authors. All authors reviewed and approved the report before submission.

## Acknowledgements

We thank the volunteers, without whom the project would not be conducted. We thank the investigators and staff of the study sites. We thank Charles A. Czeisler for his strategic advice, and colleagues at Vanda Pharmaceuticals for their trial support.

## References

1. American Academy of Sleep Medicine (AASM). International Classification of Sleep Disorders – Third Edition (ICSD-3). third edit. Darien, IL: American Academy of Sleep Medicine; 2014.

2. Weingarten JA, Collop NA. Air travel: effects of sleep deprivation and jet lag. Chest. 2013 Oct;144(1931-3543 (Electronic)):1394–401.

3. Auger RR, Burgess HJ, Emens JS, Deriy L V., Thomas SM, Sharkey KM. Clinical Practice Guideline for the Treatment of Intrinsic Circadian Rhythm Sleep-Wake Disorders: Advanced SleepWake Phase Disorder (ASWPD), Delayed Sleep-Wake Phase Disorder (DSWPD), Non-24-Hour Sleep-Wake Rhythm Disorder (N24SWD), and Irregular Sleep-Wa. J Clin Sleep Med. 2015;11(10):1199–236.

4. Lockley SW, Arendt J, Skene DJ. Visual impairment and circadian rhythm disorders. Dialogues Clin Neurosci. 2007;9 (3):301–14.

5. Waterhouse J, Reilly T, Atkinson G, Edwards B. Jet lag: trends and coping strategies. Lancet. 2007 Mar 31;369(1474-547X (Electronic)):1117–29.

6. Czeisler CA, Klerman EB. Circadian and sleep-dependent regulation of hormone release in humans. Recent ProgHormRes. 1999;54(0079-9963 (Print)):97–132.

7. Czeisler CA, Khalsa SBS. The Human Circadian Timing System and Sleep-Wake Regulation. In: Kryger MH, Roth T, Dement WC, editors. Principles and Practice of Sleep Medicine. W.B. Saunders Company; 2000. p. 353–75.

8. Berson DM, Dunn FA, Takao M. Phototransduction by retinal ganglion cells that set the circadian clock. Science. 2002 Feb 8;295(1095-9203 (Electronic)):1070–3.

9. Scheer FAJL, Hilton MF, Mantzoros CS, Shea SA. Adverse metabolic and cardiovascular consequences of circadian misalignment. Proc Natl Acad Sci [Internet]. 2009 Mar 17 [cited 2019 Feb 26];106(11):4453–8. Available from: http://www.ncbi.nlm.nih.gov/pubmed/19255424

10. Reutrakul S, Hood MM, Crowley SJ, Morgan MK, Teodori M, Knutson KL, et al. Chronotype is independently associated with glycemic control in type 2 diabetes. Diabetes Care. 2013 Sep;36_ _(9):2523–9.

11. Nguyen KD, Fentress S, Qiu Y, Yun K, Cox JS, Chawla A. Circadian Gene Bmal1 Regulates Diurnal Oscillations of Ly6Chi Inflammatory Monocytes. Science. 2013 Sep 17;341_ _(6153):1483–8.

12. Arendt J. Approaches to the Pharmacological Management of Jet Lag. Drugs [Internet]. 2018;(0123456789):1419–31. Available from: http://link.springer.com/10.1007/s40265-018-0973-8

13. Waterhouse J, Reilly T, Atkinson G. Jet-lag. Lancet. 1997 Nov 29;350(0140-6736 (Print)):1611–6. Available from: file://r/Publication PDFs/waterhouse jet lag.pdf

14. Shekleton JA, Rajaratnam SM, Gooley JJ, Van Reen E, Czeisler CA, Lockley SW. Improved neurobehavioral performance during the wake maintenance zone. J Clin Sleep Med. 2013 Apr 15;9(1550-9397 (Electronic)):353–62.

15. Rajaratnam SM, Polymeropoulos MH, Fisher DM, Roth T, Scott C, Birznieks G, et al. Melatonin agonist tasimelteon (VEC-162) for transient insomnia after sleep-time shift: two randomised controlled multicentre trials. Lancet. 2009 Feb 27;373(1474-547X (Electronic)):482–91.

16. Van Den Heuvel CJ, D.J. k, D. D. Thermoregulatory and soporific effects of very low dose melatonin injection. Am J Physiol. 1999 Feb;276_ _(2 Pt 1):E249–254.

17. Lockley SW, Dressman MA, Licamele L, Xiao C, Fisher DM, Flynn-Evans EE, et al. Tasimelteon for non-24-hour sleep-wake disorder in totally blind people (SET and RESET): two multicentre, randomised, double-masked, placebo-controlled phase 3 trials. Lancet. 2015;Epub(DOI: http://dx.doi.org/10.1016/S0140-6736(15)60031-9). Available from: doi: http://dx.doi.org/10.1016/S0140-6736(15)60031-9

18. Arendt J, Skene D. Melatonin as a chronobiotic. Sleep Med Rev. 2005;9:25–39.

19. Beaumont M, Batejat D, Pierard C, Van Beers P, Denis J, Coste O, et al. Caffeine or melatonin eff ects on sleep and sleepiness after rapid eastward transmeridian travel. J Appl Physiol. 2004;96:50–8.

20. Lewy AJ, Bauer VK, Ahmed S, Thomas KH, Cutler NL, Singer CM, et al. The human phase response curve (PRC) to melatonin is about 12 hours out of phase with the PRC to light. ChronobiolInt. 1998 Jan;15(0742–0528):71–83.

21. Samel A. Wegmann HM. Vejvoda M. Jet lag and sleepiness in aircrew. J Sleep Res. 1995;4(S2):30–6.

22. Becker T, Penzel T, Fietze I. A new German Charite Jet Lag Scale for jet lag symptoms and application. Ergonomics. 2015;58(1366-5847 (Electronic)):811–21. Available from: file://r/Publication PDFs/BECKER 2013.pdf

23. Morgenthaler TI, Lee-Chiong T, Alessi C, Friedman L, Aurora RN, Boehlecke B, et al. Practice parameters for the clinical evaluation and treatment of circadian rhythm sleep disorders. An American Academy of Sleep Medicine report. Sleep. 2007 Nov 1;30(0161-8105 (Print)):1445–59.

24. Spitzer RL, Terman M, Williams JBW, Terman JS, Malt UF, Singer F, et al. Jet lag: Clinical features, validation of a new syndrome-specific scale, and lack of response to melatonin in a randomized, double-blind trial. Am J Psychiatry. 1999;156(9):1392–6.

25. U.S.Department of Commerce Office of National Travel and Tourism ITA. TI News: An information service from the National Travel & Tourism Office (NTTO) [Internet]. 2019 [cited 2019 Apr 5]. Available from: https://travel.trade.gov/tinews/archive/tinews2019/20190402.asp

26. Travnickova-Bendova Z, Cermakian N, Reppert SM, Sassone-Corsi P. Bimodal regulation of mPeriod promoters by CREB-dependent signaling and CLOCK/BMAL1 activity. ProcNatlAcadSci USA. 2002 May 28;99(0027–8424):7728–33.

27. Srinivasan V, Singh J, Pandi-Perumal SR, Brown GM, Spence DW, Cardinali DP. Jet lag, circadian rhythm sleep disturbances, and depression: the role of melatonin and its analogs. AdvTher. 2010 Nov;27(1865-8652 (Electronic)):796–813.

28. Lavedan C, Forsberg M, Gentile AJ. Tasimelteon: A selective and unique receptor binding profile. Neuropharmacology. 2015 Jan 13;91(1873-7064 (Electronic)):142–147.

29. Zee PC, Wang-weigand S, Wright KP, Peng X, Roth T. Effects of ramelteon on insomnia symptoms induced by rapid, eastward travel. Sleep Med. 2010;11(6):525–33. Available from: http://dx.doi.org/10.1016/j.sleep.2010.03.010

30. Erland LAE, Saxena PK. Melatonin Natural Health Products and Supplements: Presence of serotonin and significant variability of melatonin content. J Clin Sleep Med. 2017;13(2):275–81.

